# EVD DURATION IN ANEURYSMAL SUBARACHNOID HAEMORRHAGE: A SINGLE CENTRE RETROSPECTIVE COHORT STUDY

**DOI:** 10.64898/2026.10.18.26365822

**Authors:** Anumeha Verma, Keryn Davidson, Emily Fitzgerald, Lachlan Donaldson, Tessa Garside, Rosalind Elliott, Frances Bass, Naomi Hammond, Alamgir Kabir, Anthony Delaney

## Abstract

**Introduction and objectives:** Hydrocephalus is a common complication of aneurysmal subarachnoid haemorrhage. This is one of several standard indications for the insertion of an external ventricular drain, but there are no standard guidelines for timing of extraventricular drain discontinuation in this population. Therefore, the study aims were to examine the 1) factors associated with extraventricular drain duration in aneurysmal subarachnoid haemorrhage and 2) association between extraventricular drain duration and clinical and resource-based outcomes.

**Methods:** In this single centre retrospective cohort study, data for adult patients treated in the ICU between 2015-2023 with aneurysmal subarachnoid haemorrhage and requiring an extraventricular drain were analysed. Data were extracted from both a prospective patient registry and the electronic medical record. Linear regression modelling was used to identify the factors associated with extraventricular drain duration. Measures used to examine the association between extraventricular drain duration, and the outcomes of interest included hazard ratio, relative risk, and mean differences as appropriate for a specific outcome.

**Results:** Of the participants (n=205), 68% (n = 140) were female, mean age was 60.7 (SD 13.3) years and 71% (n = 145) had a modified Fisher score of 4. Mean extraventricular drain duration was 15.3 (SD 8.6) days. The number of days mechanically ventilated and extraventricular drain challenge by clamp trial were associated with an increased duration of extraventricular drain treatment. Increased extraventricular drain duration was associated with an increased incidence of ventriculoperitoneal shunt insertion, and an increase in ICU and hospital length of stay when extraventricular drain duration exceeded 20 days.

**Conclusion:** The factors associated with an increased duration of extraventricular drain treatment were a longer duration of mechanical ventilation. The outcomes associated with an increased duration of extraventricular drain treatment were an increased incidence of ventriculoperitoneal shunt insertion, an increased incidence of ventriculitis when extraventricular drain duration exceeded 20 days, and an increase in ICU length of stay. These study findings may inform the conduct of a larger multi-centre study to examine the factors associated with duration of treatment with extraventricular drain, and outcomes associated with extraventricular drain duration.

**Study registration:** Not registered

## 1. Introduction

Hydrocephalus complicating aneurysmal subarachnoid haemorrhage (aSAH) occurs in up to 87% of cases of aSAH (1), and is often managed with insertion of an external ventricular drain (EVD) (1, 3–5). This allows for diversion of cerebrospinal fluid (CSF) and management of raised intracranial pressure (ICP) (1, 6). With time, the dynamics of CSF reabsorption may return to normal, allowing for EVD discontinuation (6, 7). However, weak data exist on the optimum timing of EVD discontinuation following aSAH (1, 8). This is an important aspect of care; longer EVD duration can impact on patient outcomes such as ventriculitis, an increased risk of ICU related complications, such as deep vein thrombosis (DVT) and ICU length of stay (LOS) (9, 11, 12)).

Both continuous and non-continuous CSF drainage strategies have been described in the management of hydrocephalus following aSAH (1); the EVD may be kept closed, with intermittent drainage of CSF, or open with continuous drainage (1, 4, 10). Discontinuation strategies for EVD treatment involve ‘challenging’ the EVD with a “gradual wean” or a “rapid wean” (3, 4, 10). In a gradual wean, the height of the EVD is raised every 24 hours, over 3-4 days, then the EVD is clamped (closed) (9). In a rapid wean, the EVD is closed for 24-48 hours, regardless of the height of the drain (9). In both scenarios, failure of the ‘challenge’ is defined as the patient developing symptoms (such as headache, drowsiness, or worsening neurological deficit, which may be focal), raised ICP, CSF leakage from around the EVD site, or hydrocephalus on repeat CT brain scan (9).

Some data suggest that a shorter EVD duration with an early clamp trial and intermittent drainage is safe and reduces ICU and hospital LOS (3). An intermittent drainage strategy may promote a return to normal CSF reabsorption through arachnoid granulations and may reduce the need for ventriculo-peritoneal (VP) shunt insertion (2, 10).

Surveys of practice have demonstrated many centres still undertake continuous EVD drainage with a gradual wean (2, 5, 10). A continuous EVD drainage strategy may promote the clearing of blood from SAH and reduce the risk of vasospasm and delayed cerebral ischaemia (DCI) (5, 10).

Given the paucity of evidence for the optimum EVD management and the optimum timing of EVD discontinuation in aSAH, we conducted a single-centre cohort study in patients with aSAH treated in the neurosciences ICU, with the aims of examining the:

1. factors associated with extraventricular drain duration in aneurysmal subarachnoid haemorrhage and
2. association between extraventricular drain duration and clinical and resource-based outcomes.

## 2. Methods

### 2.1 Study design, setting, participants, ethical approval, variables and data measurement

This was a single centre retrospective analysis of prospectively collected data conducted in the neurosciences adult intensive care unit, at Royal North Shore Hospital (RNSH), Australia. Adult patients (aged > 18 years) treated in RNSH ICU from the 1^st^ of January 2015 to the 31^st^ of January 2023 with a diagnosis of aSAH and with an EVD inserted were included in this study. Patients who had an EVD inserted for indications other than aSAH, or those with aSAH without an EVD were excluded. Patients with a pre-existing diagnosis of central nervous system infection were also excluded.

Approval for the study protocol and ethics approval to access the Neurological Outcomes from Intensive Care (NOICE) database and the electronic medical record (eMR) was obtained from the Human Research Ethics Committee (HREC), Northern Sydney Local Health District, prior to commencement of this study (2021/STE01272). The HREC approved a waiver such that written informed consent was not required.

Approximately 800 patients per year were admitted to the RNSH neurosciences ICU. All patients diagnosed with aSAH were initially managed within the RNSH neurosciences ICU. Baseline demographics, clinical observations, and long-term follow-up data for these patients were entered into the NOICE registry. The NOICE registry was established in RNSH in 2015. It was a prospective observational data registry that stored health information, including long-term outcomes, for all patients treated in the ICU with acute brain injuries including aSAH. Data were collected by the database manager using a specifically developed database and a predefined data dictionary. The NOICE registry was approved by the NSLHD HREC and given the low-risk nature of the study, an opt-out approach to consent was approved (HREC reference 2019/STE12306).

Baseline and aSAH characteristics were extracted from the NOICE registry from its inception from the 1^st^ of January 2015 to the 31^st^ of January 2023. Baseline characteristics included age, sex, comorbidities (ischaemic heart disease, chronic obstructive pulmonary disease, and diabetes), and illness severity, as measured by the Acute Physiology and Chronic Health Evaluation (APACHE) III score. aSAH characteristics included World Federation of Neurosurgical Societies (WFNS) grade, modified Fisher score, location of aneurysm, treatment of the aneurysm (clipping vs. coiling), and presence of vasospasm. Data regarding complications sustained during ICU admission, such as DVT were also extracted.

Further data were obtained from the eMR through review of flowsheets and clinical notes, which provided documentation of EVD use and patient management during ICU and hospital admission. Data extracted from the eMR included duration of EVD treatment (defined as the total number of days for which the EVD was in situ in the patient), timing of EVD challenge, and outcomes associated with aSAH, such as a diagnosis of ventriculitis (defined in this study by a documented diagnosis of ventriculitis by a medical officer in the eMR), or insertion of VP shunt. Other variables such as initial height of the EVD, whether the EVD was kept closed with intermittent drainage of CSF, or open with continuous drainage of CSF, were also extracted from eMR.

The method for EVD challenge occurred with either a clamp trial or height raised (3, 4, 9, 14, 15). During a clamp trial, the EVD was closed for 24-48 hours. In EVD challenge by height raised, the height of the drain was raised every 24 hours and then clamped, prior to its intended removal.

Data were collected on the outcome of the EVD challenge. A successful challenge was defined as EVD removal following the trial. If the initial EVD challenge was unsuccessful, data regarding reasons for ‘failure’ of the EVD challenge were collected. Data were also collected on EVD removal where a challenge did not occur prior to removal, or if the EVD required re-insertion.

### 2.2 Variables

The primary aim of the study was to examine the factors associated with EVD duration. The secondary aim was to examine clinical outcomes and resource use associated with EVD duration.

Factors associated with EVD duration were selected following a literature review (4, 8, 15); these included female sex, higher grade aSAH, posterior circulation aneurysms and the technique by which EVD “challenge” occurred (4, 8, 15). Factors that had a physiological basis for inclusion were also included, such as age of the patient, presence of co-morbidities, duration of mechanical ventilation, and whether there was a daily documented plan for EVD management by the neurosurgeons.

The primary outcome measure was the duration of EVD treatment, which was defined as the number of days the EVD was in situ in the patient. This was sourced from the NOICE registry. Secondary outcome measures included clinical and resource-based outcomes; clinical outcomes included the incidence of DVT, ventriculitis, VP shunt insertion, and mortality. All were sourced from eMR. Other secondary clinical outcomes included long-term functional outcomes at 6- and 12-months post-ictus; these were obtained from the NOICE registry and were measured using the Extended Glasgow Outcome Scale (GOSE).

The GOSE score was determined via a structured questionnaire conducted during a follow-up phone call at 6- and 12-month post ictus. The questionnaire used pre-determined criteria and was available through the NOICE registry resources. In this study, GOSE scores of 2-4 indicated ‘severe disability’, 5-6 indicated ‘moderate disability’ and 7-8 indicated ‘recovery’. Resource-based secondary outcomes included the number of days ventilated, and ICU and hospital LOS, and were also extracted from the NOICE registry.

### 2.3 Statistical methods

Descriptive statistics were used to summarise the baseline and EVD management characteristics. The frequency and percentage of all the categories of the characteristic variables were presented. The mean and standard deviation (SD) of EVD duration were calculated for each category of the independent variables, to provide comparative data to other similar studies (3, 14, 15, 17). Univariate and multivariable linear regression models were used to estimate the unadjusted and adjusted regression coefficients for each variable, along with their corresponding 95% confidence intervals (CIs).

To address the secondary aims, EVD duration was categorised into quartiles to examine its association with secondary outcomes. For binary outcomes, the number and percentage of outcomes were reported for each quartile. Relative risks (RRs) and 95% CIs were estimated using univariate and multivariable Poisson regression models, with the first quartile serving as the reference group. The multivariable model adjusted for age, sex, number of days ventilated, type of EVD challenge, and WFNS score. For analyses of the secondary outcomes, VP shunt insertion and ventriculitis, we incorporated an offset term for person-time at risk. Follow-up time was censored at death occurring within 6 months to appropriately account for differential time contributed to the study. Hazard ratios were calculated as part of the analysis for mortality at 6 months follow-up, with the first quartile serving as the reference group. Hazard ratios were adjusted for age, sex, number of days ventilated, and type of EVD challenge.

For continuous secondary outcomes, such as ICU LOS and hospital LOS, the mean (SD) was reported for each EVD duration quartile. Association between EVD duration quartile and these continuous outcomes were assessed using univariate and multivariable linear regression models, with β-coefficients and 95% CIs reported. The multivariable models included the same covariates as used in the Poisson regression models to adjust the β-coefficients.

## 3. Results

### 3.1 Participant and aSAH characteristics

Between the 1^st^ of January 2015 and the 31^st^ of January 2023, 545 patients were treated in the RNSH neurosciences ICU for a diagnosis of aSAH and screened for inclusion. Of these, 205 patients with a diagnosis of aSAH requiring an EVD were included in the study (Figure 1). The mean (SD) duration of EVD treatment was 15.3 (8.6) days.

**Figure 1.**
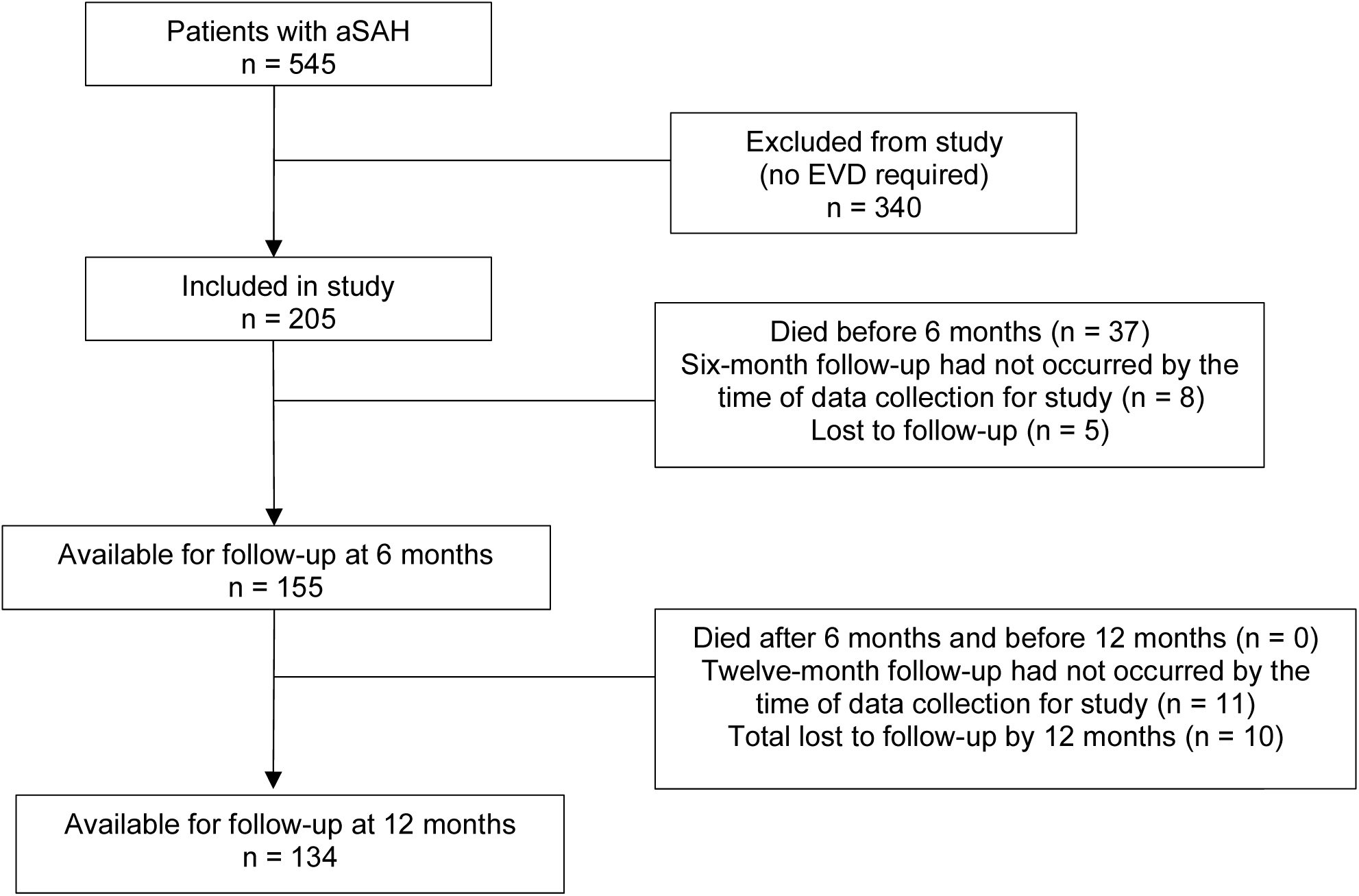
Flow of participants through the study aSAH = aneurysmal subarachnoid haemorrhage EVD = External ventricular drain

The mean (SD) age of the cohort was 60.7 (13.3) years and 68.6% were female (Table 1). A continuous drainage strategy for EVD treatment was used for 94% of the cohort (Table 2). EVD challenge began at a mean (SD) of 14.27 (6.9) days following insertion of EVD. There were 58 participants that did not undergo EVD challenge, therefore EVD challenge occurred in 118/(205–58) (80.3%) with a clamp trial.

**Table 1.**
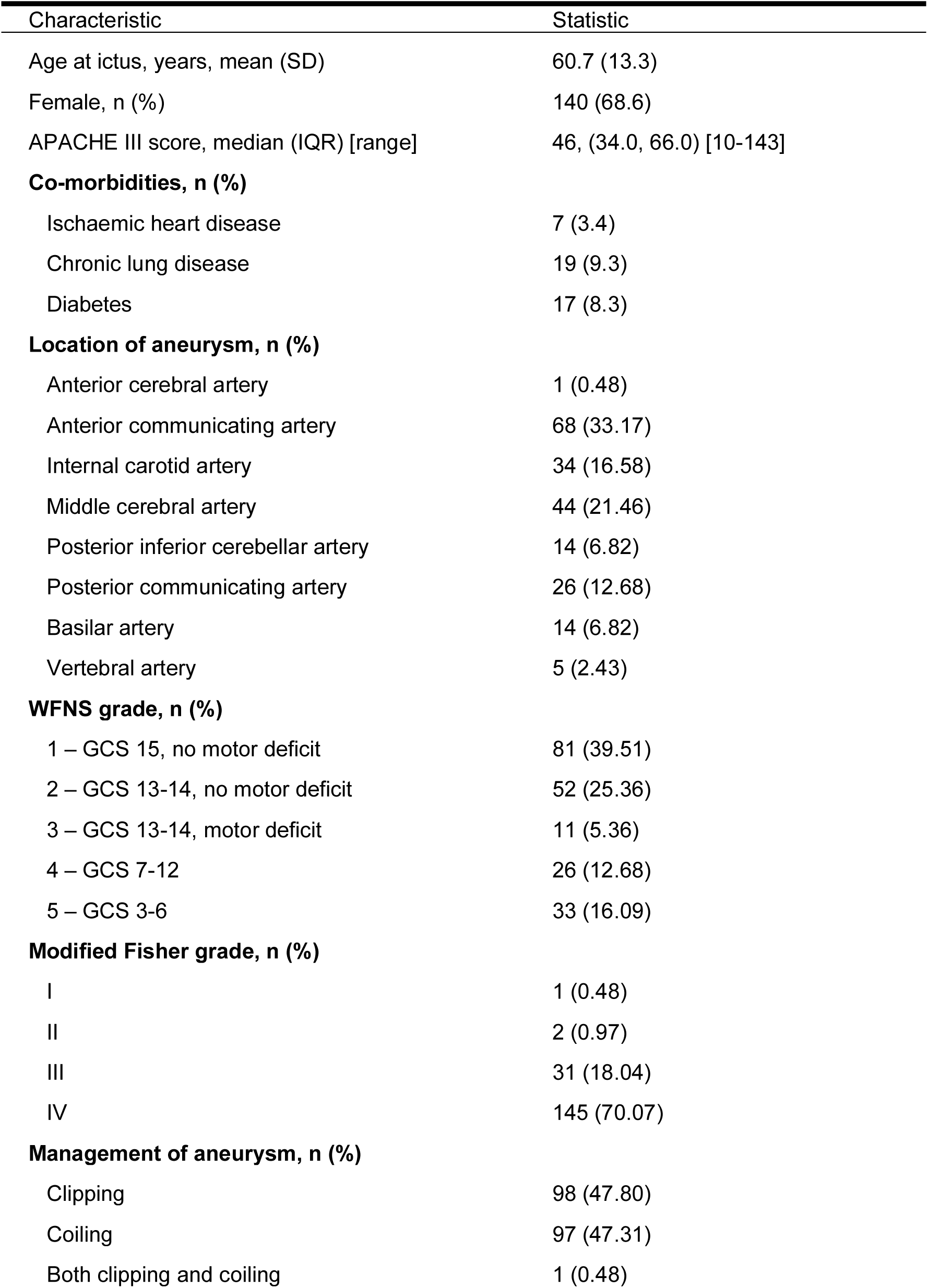
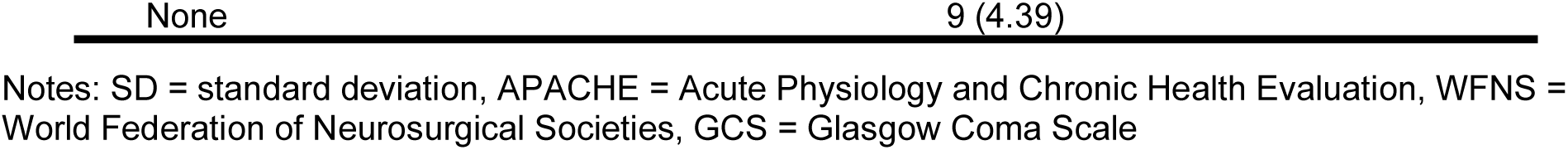
Baseline characteristics of patients and aneurysmal subarachnoid haemorrhage (n =205)

| Characteristic | Statistic |
| --- | --- |
| Age at ictus, years, mean (SD) | 60.7 (13.3) |
| Female, n (%) | 140 (68.6) |
| APACHE III score, median (IQR) [range] | 46, (34.0, 66.0) [10-143] |
| **Co-morbidities, n (%)** |  |
| Ischaemic heart disease | 7 (3.4) |
| Chronic lung disease | 19 (9.3) |
| Diabetes | 17 (8.3) |
| **Location of aneurysm, n (%)** |  |
| Anterior cerebral artery | 1 (0.48) |
| Anterior communicating artery | 68 (33.17) |
| Internal carotid artery | 34 (16.58) |
| Middle cerebral artery | 44 (21.46) |
| Posterior inferior cerebellar artery | 14 (6.82) |
| Posterior communicating artery | 26 (12.68) |
| Basilar artery | 14 (6.82) |
| Vertebral artery | 5 (2.43) |
| **WFNS grade, n (%)** |  |
| 1 – GCS 15, no motor deficit | 81 (39.51) |
| 2 – GCS 13-14, no motor deficit | 52 (25.36) |
| 3 – GCS 13-14, motor deficit | 11 (5.36) |
| 4 – GCS 7-12 | 26 (12.68) |
| 5 – GCS 3-6 | 33 (16.09) |
| **Modified Fisher grade, n (%)** |  |
| I | 1 (0.48) |
| II | 2 (0.97) |
| III | 31 (18.04) |
| IV | 145 (70.07) |
| **Management of aneurysm, n (%)** |  |
| Clipping | 98 (47.80) |
| Coiling | 97 (47.31) |
| Both clipping and coiling | 1 (0.48) |

Notes: SD = standard deviation, APACHE = Acute Physiology and Chronic Health Evaluation, WFNS = World Federation of Neurosurgical Societies, GCS = Glasgow Coma Scale

**Table 2.** Characteristics of extraventricular drain management (n = 205)

| Characteristics | Statistic |
| --- | --- |
| EVD duration, days, mean (SD) | 15.4 (8.6) |
| Continuous drainage, n (%) | 193 (94.1) |
| Days until EVD challenge post EVD insertion, mean (SD) | 14.27 (6.9) |
| EVD clamp trial, n (%) | 118 (57.6) |
| EVD height raised, n (%) | 16 (7.8) |
| Reason EVD not challenged, n (%) | 58 (28.3) |
| Withdrawal of life-sustaining treatment | 22 |
| Patient died with EVD in situ | 7 |
| EVD blocked and removed | 11 |
| EVD dislodged | 13 |
| Clamped from insertion | 2 |
| Unspecified | 3 |
| Reason EVD failed initial challenge, n (%) | 33 (16) |
| Raised ICP | 4 |
| Ventriculomegaly on CTB | 9 |
| CSF leak at exit site | 8 |
| Clinical deterioration | 8 |
| Unspecified | 4 |
| Daily EVD management plan documented in clinical record, n (%) | 6 (2.9) |

Notes: EVD = external ventricular drain, SD = standard deviation

### 3.2 Factors associated with EVD duration

The factors associated with EVD duration were outlined in Table 3. Patients in the third quartile of the APACHE score experienced a significantly shorter mean EVD duration compared to those in the lowest quartile (adjusted β: −5.37; 95% CI: −9.16 to −1.58). An association was evident between EVD duration, and the number of days patients were ventilated. Specifically, compared to the first quartile, EVD duration increased by an average of 2.22 days in the second quartile (Adj. β: 2.22; 95% CI: −1.26 to 5.70), 7.55 days in the third quartile (Adj β: 7.55; 95% CI: 4.38 to 10.71), and 13.08 days in the fourth quartile (Adj β: 13.08; 95% CI: 9.71 to 16.44).

**Table 3.**
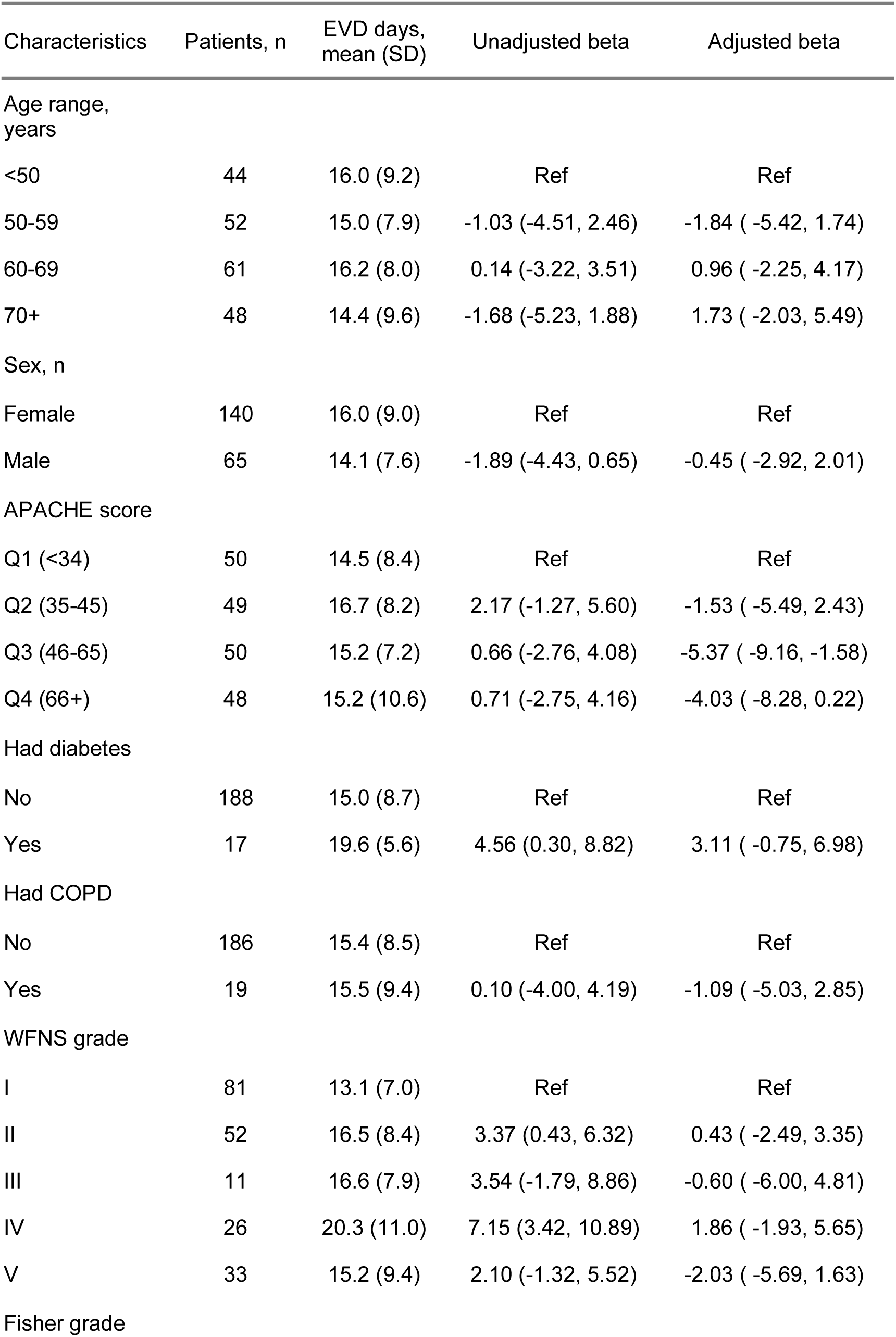
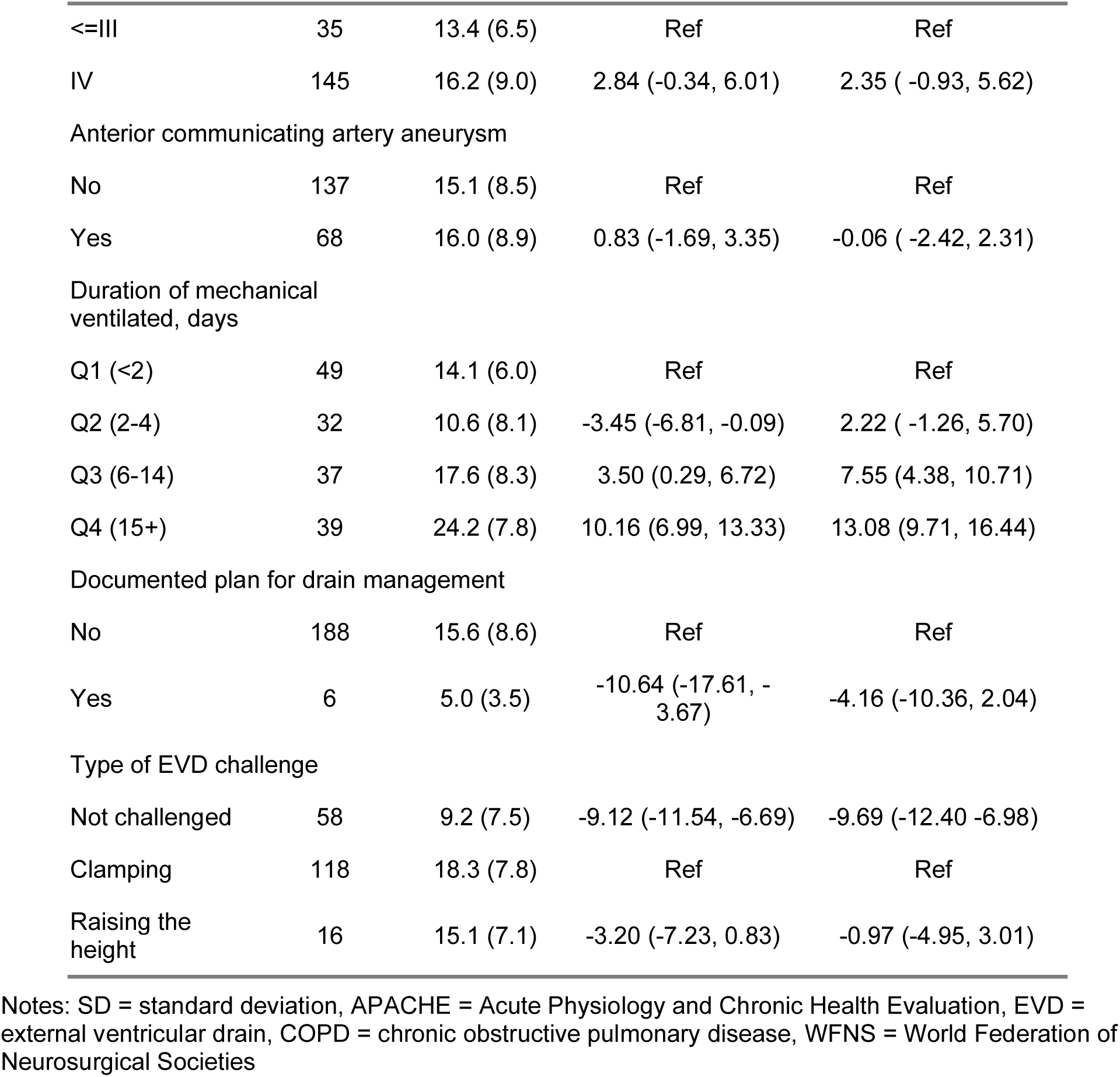
Factors associated with extraventricular drain duration.

Patients who did not have an EVD challenge had significantly shorter EVD durations than those who underwent EVD challenge by clamping (Adj. β: −9.69; 95% CI: − −12.40 to −6.98). EVD challenge by height raised was associated with shorter EVD duration by 3 days than challenge by clamping, but this was not statistically significant (Adj. β: −0.97; 95% CI: − −4.95 to −3.01).

### 3.3 Clinical outcomes associated with EVD duration

The clinical outcomes associated with EVD duration were outlined in **Tables 4** and **5** and **Supplementary Tables 1.1-2.6**. An increased duration of EVD treatment was associated with an increased likelihood of VP shunt insertion. Compared to the patients in quartile 1 of EVD duration, quartile 4 was associated with a 6.03-fold (Adj. RR: 6.03; 95% CI: 1.45 to 32.56) increased risk of VP shunt insertion (**Table 4**).

**Table 4:** Association between extraventricular drain duration and insertion of ventriculoperitoneal shunt.

|  | No | Yes | Crude RR<sup>1</sup> (95% CI) | Adj RR<sup>1</sup> (95% CI) | p-value |
| --- | --- | --- | --- | --- | --- |
| EVD duration<br>(days) |  |  |  |  |  |
| Quartile-1<br>(1.5 - 9.0) | 52 (91%) | 5 (8.8%) | 1 | 1 |  |
| Quartile-2<br>(9.01 – 15.8) | 40 (87%) | 6 (13%) | 1.37 (0.41, 4.77) | 1.04 (0.20, 6.00) | 0.960 |
| Quartile-3<br>(15.81 – 20.0) | 40 (74%) | 14 (26%) | 2.60 (1.00, 8.06) | 2.31 (0.59, 11.86) | 0.260 |
| Quartile-4<br>(20.01 – 52.46) | 19 (40%) | 29 (60%) | 6.01 (2.52, 17.68) | 6.03 (1.45, 32.56) | 0.021 |

<sup>1</sup>Relative risks were estimated using Poisson model with an offset for person-time at risk; follow-up was censored at death within 6 months to account for unequal observation time.
Notes: RR = relative risk, EVD = external ventricular drain, CI = confidence interval

**Table 5:** Association between extraventricular drain duration and ventriculitis.

|  | No | Yes | Crude RR<sup>1</sup> (95% CI) | Adj RR<sup>1</sup> (95% CI) | p-value |
| --- | --- | --- | --- | --- | --- |
| EVD duration<br>(days) |  |  |  |  |  |
| Quartile-1<br>(1.5 - 9.0) | 50 (88%) | 7 (12%) | 1 | 1 |  |
| Quartile-2 (9.01 – 15.8) | 34 (74%) | 12 (26%) | 1.96 (0.79, 5.27) | 2.22 (0.46, 10.57) | 0.314 |
| Quartile-3 (15.81 – 20.0) | 33 (61%) | 21 (39%) | 2.79 (1.25, 7.09) | 2.90 (0.61, 13.83) | 0.181 |
| Quartile-4 (20.01 – 52.46) | 31 (65%) | 17 (35%) | 2.230 (0.97, 6.02) | 4.91 (0.900, 26.800) | 0.066 |

<sup>1</sup>Relative risks were estimated using Poisson model with an offset for person-time at risk; follow-up was censored at death within 6 months to account for unequal observation time.
Notes: RR = relative risk, EVD = external ventricular drain, CI = confidence interval,

Longer EVD duration was associated with an increased risk of ventriculitis, although the association was not statistically significant. The incidence of ventriculitis for EVD durations over 20 days was significantly higher when compared to <9 days in the adjusted analysis (**Table 5**). The adjusted relative risk of DVT was higher with an increased duration of EVD (**Supplementary Table 2.1)**, however this was not statistically significant.

An increased duration of EVD treatment was associated with an increase in ICU LOS **(Supplementary Table 2.2).** An increased duration of EVD treatment was associated with an increased hospital LOS when EVD treatment exceeded 20 days **(Supplementary Table 2.3).** While there was a trend towards increased risk of severe disability with increased EVD duration, this result did not reach statistical significance **(Supplementary tables 2.4-2.5)**. An association was observed between EVD duration and mortality at 6 months, as the hazard ratio reduced with an increase in EVD duration, however EVD duration was not associated with a statistically significant change in mortality **(Supplementary table 2.6)**.

## 4. Discussion

### 4.1 Key findings

This was a single centre retrospective cohort study. The aim was to examine the factors associated with duration of EVD treatment, and to identify the outcomes associated with duration of EVD treatment. The factors associated with an increased duration of EVD treatment included an EVD challenge by clamp trial, and the number of days ventilated.

The clinical outcomes associated with an increased duration of EVD treatment were an increased incidence of VP shunt insertion, and an increase in ICU LOS. Longer duration of EVD treatment was associated with an increased risk of ventriculitis when EVD duration exceeded 20 days,

### 4.2 Interpretation and generalisability

The duration of EVD treatment in our centre was longer than in other centres. In our centre, discontinuation of EVD treatment occurred at a mean of 15.3 days. In other centres mean EVD duration ranged from 5-15.2 days (2, 3, 14, 18, 19). The timing of EVD challenge in our study occurred at a mean of 14.27 days EVD duration and mostly occurred with clamp trial. In other centres, EVD challenge began as early as 3-5 days post-insertion (18), or when the peak vasospasm period had resolved (beginning 7 days post-ictus) (2, 17). The timing and method of EVD challenge were at the discretion of the neurosurgeons at our facility, like other centres (4, 8, 17).

We found an increase in EVD duration with challenge by clamp trial. This contrasted with other studies, which reported an increase in EVD duration with gradual EVD wean (2, 3, 14). This was likely irrelevant, as the sample size for patients that underwent EVD challenge with a gradual wean in our study was small. Patients that did not undergo EVD challenge included patients that had short EVD duration due to death, blockage and removal of the EVD, or dislodgment of the EVD.

Other factors that had been found to be associated with EVD duration in other studies, such as female sex, higher grade aSAH and posterior circulation aneurysms (4, 8, 15) were not associated with EVD duration in our study. The association between higher grade aSAH and EVD duration may have physiological basis, as increased EVD duration may assist with the clearing of blood from the ventricles (5, 10).

Increased EVD duration was associated with an increased incidence of VP shunt, consistent with other centres with similar cohorts (14, 17, 19). The incidence of VP shunt in our study fell within the range described at other centres with similar cohorts, from 6-67% (17, 18).

We found an increased EVD duration was associated with an increased risk of ventriculitis, but when adjusted for other variables, these results were not statistically significant. However, the incidence of ventriculitis for EVD durations over 20 days was significantly higher when compared to <9 days in the adjusted analysis in our study.

Several studies identified that the most consistent variable associated with ventriculitis was EVD duration (11, 13, 16, 20, 21). Other predictors of ventriculitis included the use of multiple EVDs, EVD manipulation frequency, non-adherence to strict EVD maintenance protocols, and systemic infections (11, 16). In some centres, the incidence of ventriculitis decreased after 11 days post-EVD insertion, suggesting that infection was introduced at the time of EVD insertion, and therefore resolved to use a single ICP monitor for as long as possible and necessary(11).

The rate of ventriculitis in our cohort fell within the 5-35% range of EVD-related ventriculitis described in other centres (13, 22); however, that range accounted for the use of EVDs in aSAH as well as other indications such as hypertensive intracerebral bleeds. Additionally, the definition of ventriculitis used in these centres was not clarified (13, 22). The protocol at our centre typically involved the use of a single ICP monitor. Replacement of the EVD occurred only if there was malfunction of the existing EVD and there was ongoing need for an EVD as determined by the neurosurgeon. Surveillance cultures were taken from the EVD twice a week, to reduce the risk of ventriculitis from more frequent handling of the EVD.

The association between EVD duration and duration of mechanical ventilation was an expected finding, as an EVD would likely have remained in situ as a neurological monitoring tool in a comatose patient.

The association between increased EVD duration and reduced mortality was an unexpected finding. This association may be irrelevant, as EVD treatment would be discontinued if participants died, and they likely died due to reasons other than the duration of EVD treatment. Further investigation is warranted to determine the association between mortality in the study population and other factors including age, APACHE score, co-morbidities and WFNS grade. The association between EVD duration and ICU LOS and hospital LOS was consistent with other studies (3, 4, 13, 14).

### 4.3 Strengths and limitations

This study was one of the larger single centre studies to examine the factors associated with EVD duration in aSAH and the outcomes associated with EVD duration. Over eight years of data were available and were comparable to other similar studies (3, 14, 17).

The study was limited by its retrospective design, though the data in the NOICE registry had been prospectively collected. This study was underpowered due to the small number of patients. The study population was from a single hospital, limiting external validity.

We had a small sample size for patients that had EVD challenge with a gradual wean approach, which may have affected the findings. Data regarding the timespan that the ‘height raised’ challenge occurred over would also have been beneficial and may have influenced the findings. Many participants who did not have their EVD challenged had died; the duration of the EVD in this group was confounded by mortality. This could be accounted for in a competing risks analysis. In the analysis of outcomes associated with EVD duration, the competing risk of death may potentially have impacted on all the other outcomes in the analysis, including the incidence of VP shunt, ventriculitis and DVT, ICU LOS and hospital LOS. These outcomes may have been further investigated with an analysis of the competing risk of mortality.

The diagnosis of ventriculitis is challenging and several definitions have been described (20). In our study, ventriculitis was defined by a documented diagnosis of ventriculitis in eMR by the intensive care medical team. The subjective nature of the definition of ventriculitis may have influenced our findings. Given the retrospective nature of the study, accounting for variations in the definition of ventriculitis amongst the physicians involved in the care of the patients was difficult to control for. This was a limitation of the study.

Other limitations included the unavailability of electronic medical records prior to 2018, loss to follow-up and missing data in the NOICE database. For data prior to 2018, we could only obtain the initial height of the EVD, and not trends regarding EVD output, height, median daily ICP, duration of vasospasm and its treatment duration. Our analysis was restricted to the variables available in the NOICE database, potentially omitting other important factors that could affect the validity of the study, such as daily EVD output, daily maximum ICP, vasospasm and its treatment. Consequently, our findings may not be generalizable to the target population. Further studies with a larger sample size and a broader range of covariates are needed to confirm the findings of this study.

## 5. Conclusions

The factors associated with an increased duration of EVD treatment were EVD challenge were a longer duration of ventilation, which was an intuitive association. The outcomes associated with an increased duration of EVD treatment were an increased incidence of VP shunt insertion, an increased risk of ventriculitis when EVD duration exceeded 20 days, and an increase in ICU LOS. This study provides the observational data to inform the conduct of a larger multi-centre study to examine the factors associated with duration of treatment with EVD, and outcomes associated with EVD duration.

## Author contributions (CRediT statement)

Anumeha Verma: Conceptualisation, investigation, writing (original draft, review, editing), visualisation

Keryn Davidson: Conceptualisation, writing – review and editing

Emily Fitzgerald: Resources, data curation, writing – review and editing

Lachlan Donaldson: Writing – review and editing

Tessa Garside: Writing – review and editing

Rosalind Elliott: Writing – review and editing

Frances Bass: Project administration, writing – review and editing

Naomi Hammond: Resources, writing – review and editing

Alamgir Kabir: Data curation, formal analysis

Anthony Delaney: Conceptualisation, methodology, supervision, writing – review and editing

## Data Availability

All data produced in the present study are available upon reasonable request to the authors.

Due to the nature of this research, participants of this study did not agree for their data to be shared publicly, so supporting data is not available.

## Acknowledgments

The authors acknowledge the contribution of all the study participants and their families. We would also like to thank those who assisted in the collection of data for the NOICE registry.

