## Supplementary tables for "EVD DURATION IN ANEURYSMAL SUBARACHNOID HAEMORRHAGE: A SINGLE CENTRE RETROSPECTIVE COHORT STUDY"

### Supplementary Appendix

Supplementary results:

**Supplementary Table 1.1** EVD duration by primary and secondary outcome categories

| Outcomes | n | Mean (SD) |
|---|---|---|
| Overall | 205 | 15.4 (8.6) |
| Mortality at 6 months | | |
| Alive | 154 | 16.8 (8.2) |
| Died | 39 | 10.2 (8.6) |
| Functional outcome at 6 months | | |
| Recovery | 56 | 14.1 (7.0) |
| Moderate disability | 43 | 15.4 (6.5) |
| Severe disability | 53 | 20.5 (9.2) |
| Functional outcome at 12 months | | |
| Recovery | 59 | 14.3 (7.0) |
| Moderate disability | 34 | 16.7 (6.6) |
| Severe disability | 49 | 20.6 (8.7) |
| Insertion VP Shunt | | |
| No | 151 | 13.0 (7.0) |
| Yes | 54 | 22.2 (9.2) |
| Diagnosis of ventriculitis | | |
| No | 148 | 14.1 (8.1) |
| Yes | 57 | 19.0 (8.9) |
| ICU hours | | |
| Quartile1 (1.5 - 9.0 days) | 52 | 6.2 (3.8) |
| Quartile2 (9.01 – 15.8 days) | 51 | 13.3 (4.4) |
| Quartile3 (15.81 – 20.0 days) | 51 | 17.2 (3.8) |
| Quartile4 (20.01 – 52.46 days) | 51 | 25.2 (7.7) |
| Hospital hours | | |
| Quartile1 (1.5 - 9.0 days) | 51 | 7.6 (5.0) |

| Outcomes | n | Mean (SD) |
|---|---|---|
| Quartile2 (9.01 – 15.8 days) | 51 | 13.5 (5.4) |
| Quartile3 (15.81 – 20.0 days) | 50 | 17.2 (5.9) |
| Quartile4 (20.01 – 52.46 days) | 51 | 23.0 (9.0) |

**Supplementary table 2.1:** Association between extraventricular drain duration and deep vein thrombosis

| | No | Yes | Crude RR (95% CI) | Adj RR (95% CI) | p-value |
|---|---|---|---|---|---|
| EVD duration (days) | | | | | |
| Quartile-1 (1.5 - 9.0) | 24 (89%) | 3 (11%) | 1 | 1 | |
| Quartile-2 (9.01 – 15.8) | 35 (92%) | 3 (7.9%) | 0.71 (0.13, 3.84) | 0.71 (0.07, 6.97) | 0.800 |
| Quartile-3 (15.81 – 20.0) | 41 (89%) | 5 (11%) | 0.98 (0.24, 4.77) | 1.67 (0.27, 14.3) | 0.600 |
| Quartile-4 (20.01 – 52.46) | 29 (62%) | 18 (38%) | 3.45 (1.17, 14.7) | 2.81 (0.41, 25.8) | 0.300 |

Notes: RR = relative risk, EVD = external ventricular drain, CI = confidence interval

**Supplementary table 2.2:** Association between extraventricular drain duration and ICU length of stay

| | n | Mean (SD) | Crude β (95% CI) | Adj. β (95% CI) | p-value |
|---|---|---|---|---|---|
| EVD duration (days) | | | | | |
| Quartile-1 (1.5 - 9.0) | 57 | 10.2 (6.2) | Ref | Ref | |
| Quartile-2 (9.01 – 15.8) | 46 | 16.4 (4.4) | 6.2 (4.1, 8.2) | 5.7 (2.7, 8.8) | <0.001 |
| Quartile-3 (15.81 – 20.0) | 54 | 20.6 (2.8) | 10.4 (8.4, 12) | 9.7 (6.5, 13) | <0.001 |
| Quartile-4 (20.01 – 52.46) | 48 | 29.4 (6.5) | 19.2 (17, 21) | 17.0 (13, 20) | <0.001 |

Notes: SD = standard deviation, RR = relative risk, EVD = external ventricular drain, CI = confidence interval

**Supplementary table 2.3:** Association between extraventricular drain duration and hospital length of stay (days)

| | n | Mean (SD) | Crude β (95% CI) | Adj. β (95% CI) | p-value |
|---|---|---|---|---|---|
| EVD duration (days) | | | | | |
| Quartile-1 (1.5 - 9.0) | 57 | 18.8 (19.3) | Ref | Ref | |
| Quartile-2 (9.01 – 15.8) | 46 | 25.8 (9.8) | 7.0 (-1.3, 15.0) | 1.2 (-11.0, 14.0) | 0.900 |
| Quartile-3 (15.81 – 20.0) | 53 | 34.7 (19.0) | 16.0 (7.9, 24.0) | 10.0 (-3.1, 23.0) | 0.140 |
| Quartile-4 (20.01 – 52.46) | 47 | 48.9 (31.9) | 30.1 (22.0, 38.0) | 21.0 (6.3, 36.0) | 0.006 |

Notes: SD = standard deviation, RR = relative risk, EVD = external ventricular drain, CI = confidence interval

**Supplementary table 2.4:** Association between EVD duration and functional outcome (good recovery-GOSE scale = 7 and 8) at 6 months follow up

| | Did not have good recovery | Had good recovery (GOSE-7 & 8) | Crude RR (95% CI) | Crude p-value | Adj RR (95% CI) | Adj. p-value |
|---|---|---|---|---|---|---|
| EVD duration (days) | | | | | | |
| Quartile-1 (1.5 - 9.0) | 16 (48%) | 17 (52%) | 1 | | 1 | |
| Quartile-2 (9.01 – 15.8) | 19 (58%) | 14 (42%) | 0.82 (0.40, 1.67) | 0.600 | 1.65 (0.38, 8.14) | 0.500 |
| Quartile-3 (15.81 – 20.0) | 32 (70%) | 14 (30%) | 0.59 (0.29, 1.20) | 0.140 | 1.19 (0.29, 5.39) | 0.800 |
| Quartile-4 (20.01 – 52.46) | 29 (73%) | 11 (28%) | 0.53 (0.24, 1.13) | 0.100 | 2.59 (0.60, 12.7) | 0.200 |

Relative Risks (RRs) were adjusted for age, sex, number of days ventilated, and Type of EVD challenge

**Supplementary Table 2.5:** Association between EVD duration and functional outcome (good recovery- GOSE scale = 7 and 8) at 12 months follow up

| | Did not have good recovery | Had good recovery (GOSE-7 & 8) | Crude RR (95% CI) | Crude p-value | Adj RR (95% CI) | Adj. p-value |
|---|---|---|---|---|---|---|
| EVD duration (days) | | | | | | |
| Quartile-1 (1.5 - 9.0) | 16 (48%) | 17 (52%) | 1 | | 1 | |
| Quartile-2 (9.01 – 15.8) | 19 (58%) | 14 (42%) | 0.82 (0.40, 1.67) | 0.600 | 1.65 (0.38, 8.14) | 0.500 |
| Quartile-3 (15.81 – 20.0) | 32 (70%) | 14 (30%) | 0.59 (0.29, 1.20) | 0.140 | 1.19 (0.29, 5.39) | 0.800 |
| Quartile-4 (20.01 – 52.46) | 29 (73%) | 11 (28%) | 0.53 (0.24, 1.13) | 0.100 | 2.59 (0.60, 12.7) | 0.200 |

RRs were adjusted for age, sex, number of days ventilated, and Type of EVD challenge

**Supplementary Table 2.6:** Association between EVD duration and mortality at 6 months follow up

| | **Alive**, N = 154 | Died | Crude HR (95% CI) | Crude p-value | Adj HR (95% CI) | Adj. p-value |
|---|---|---|---|---|---|---|
| EVD duration (days) | | | | | | |
| Quartile-1 (1.5 - 9.0) | 33 (60%) | 22 (40%) | 1 | | 1 | |
| Quartile-2 (9.01 – 15.8) | 33 (80%) | 8 (20%) | 0.37 (0.16, 0.86) | 0.021 | 0.76 (0.21, 2.79) | 0.700 |
| Quartile-3 (15.81 – 20.0) | 46 (90%) | 5 (9.8%) | 0.19 (0.07, 0.51) | <0.001 | 0.34 (0.08, 1.49) | 0.200 |
| Quartile-4 (20.01 – 52.46) | 42 (91%) | 4 (8.7%) | 0.09 (0.02, 0.37) | <0.001 | 0.08 (0.01, 0.53) | 0.009 |

Hazard Ratios (HRs) were adjusted for age, sex, number of days ventilated, and Type of EVD challenge
